# Temporal Association Between Particulate Matter Pollution and Case Fatality Rate of COVID-19 in Wuhan, China

**DOI:** 10.1101/2020.04.09.20049924

**Authors:** Ye Yao, Jinhua Pan, Zhixi Liu, Xia Meng, Weidong Wang, Haidong Kan, Weibing Wang

## Abstract

The Coronavirus (COVID-19) epidemic, which was first reported in December 2019 in Wuhan, China, has caused 3,314 death as of March 31, 2020 in China. This study aimed to investigate the temporal association between case fatality rate (CFR) of COVID-19 and particulate matter (PM) in Wuhan. We conducted a time series analysis to explore the temporal day-by-day associations. We found COVID-19 held higher case fatality rate with increasing concentrations of PM_2.5_ and PM_10_ in temporal scale, which may affect the process of patients developed from mild to severe and finally influence the prognosis of COVID-19 patients.

## Introduction

COVID-19 is a new emerging infectious disease that poses massive challenges to global health and the economy. As of March 31, 2020, there have been more than 82,601 confirmed cases in China, and a total of 3,314 deaths have occurred in China. Wuhan city, the source of the outbreak, have accounted for 61.0% of the total number of cases and 76.9% of the deaths in China. Some researchers have found that air pollution can affect the case fatality rate (CFR) of severe acute respiratory syndrome (SARS)^1^. The COVID-19, as a respiratory disease which has a certain degree of similarity with SARS, may also have some relations between CFR and air pollution^2^. Particulate matter (PM) is the main primary air pollutant. Therefore, this study aimed to investigate the temporal association between CFR of COVID-19 and PM in Wuhan.

## Methods

We collected COVID-19 confirmed cases and deaths information which was reported by the National Health Commission. We defined case fatality rate (CFR) as deaths at day.x /new infection cases at day.x-^3^ (where T=average time period (T) from case infection to death^1^). Daily CFR were calculated for Wuhan city from January 19 to March 15 (very few confirmed cases afterwards). We applied several ways to estimate the average time period from case infection to death. Firstly, the median time from illness to death of a large sample in China was reported as 18.9 days^4^. Secondly, we found that the peak time of new diagnosis cases in Wuhan should be around February 5 and the peak time of new deaths in Wuhan is February 23, with a difference of 18 days. Adding the 4-day average time from infection to confirmation^4^, the time period should be around 22 days. Thirdly, as reported by Chinese CDC, most deaths happened 2 weeks to 8 weeks after patients’ infections (http://www.nhc.gov.cn/jkj/s3578/202002/87fd92510d094e4b9bad597608f5cc2c.shtml). Thus, we believed that the average time period should be around 21 days, which was consistent with some other study based on a large cohort study^5^. Then, we assumed the average time period from case infection to death is 21 days and calculated CFR with a 21-day lag in this study. In addition, we checked the results based on the lag time varying from 19 to 23 days, and reached the same conclusion.

We also collected daily fine particulate matter (PM_2.5_), and inhalable particulate matter (PM_10_) from National Urban Air Quality Publishing Platform (http://106.37.208.233:20035/), and meteorological data including daily mean temperature and relative humidity from the China Meteorological Data Sharing Service System. We conducted a time series analysis to explore the temporal day-by-day associations of PM_2.5_ and PM_10_ with CFR of COVID-19. We also examined the lag effects and patterns of PM_2.5_ and PM_10_ on CFR by analyzing the associations between CFR of COVID-19 and single-day PM levels on current day (lag0) and up to 5 days (lag1 – lag5) before the date of infections.

## Results

Between 19 January 2020 to 15 March 2020, the daily CFR averaged 6.4 with a range of 1.5%-13.2%, while mean daily PM_2.5_ and PM_10_ were 47.3 and 56.1 respectively (range: 10.7-100.0 μg/m^3^; 20.3-112.6 μg/m^3^) (**Table 1**). The temporal trend of daily CFR is highly similar to the temporal variation curves of PM_2.5_ and PM_10_ concentrations (**Figure 1**).

**Table 1.** Description of Daily Case Counts and Weather Conditions.

|  | Mean | SD | Min. | Median | Max. |
| --- | --- | --- | --- | --- | --- |
| Cases |  |  |  |  |  |
| Infection Case Number | 788.76 | 489.89 | 94 | 779 | 1516 |
| Death Count | 50.32 | 35.23 | 6 | 37 | 131 |
| Case Fatality Rate (%) | 6.43 | 2.97 | 1.51 | 5.94 | 13.24 |
| Meteorological Variables and Air Pollutants |  |  |  |  |  |
| Mean Temperature (°C) | 7.18 | 4.02 | 1.80 | 5.85 | 18.70 |
| Maximum Temperature (°C) | 12.75 | 4.78 | 4.70 | 12.85 | 24.90 |
| Minimum Temperature (°C) | 3.02 | 4.44 | -2.70 | 2.85 | 14.50 |
| Relative Humidity (%) | 81.37 | 8.35 | 59.00 | 82.00 | 93.00 |
| PM<sub>2.5</sub> (µg/m<sup>3</sup>) | 47.25 | 24.16 | 10.71 | 41.77 | 99.96 |
| PM<sub>10</sub> (µg/m<sup>3</sup>) | 56.07 | 26.50 | 20.33 | 52.77 | 112.58 |

\* Death counts and case fatality rate were from 8 Feb. to 15 Mar. while all the other indexes were from 19 Jan. to 25 Feb, as demonstrated in Figure 1.

**Figure 1.**
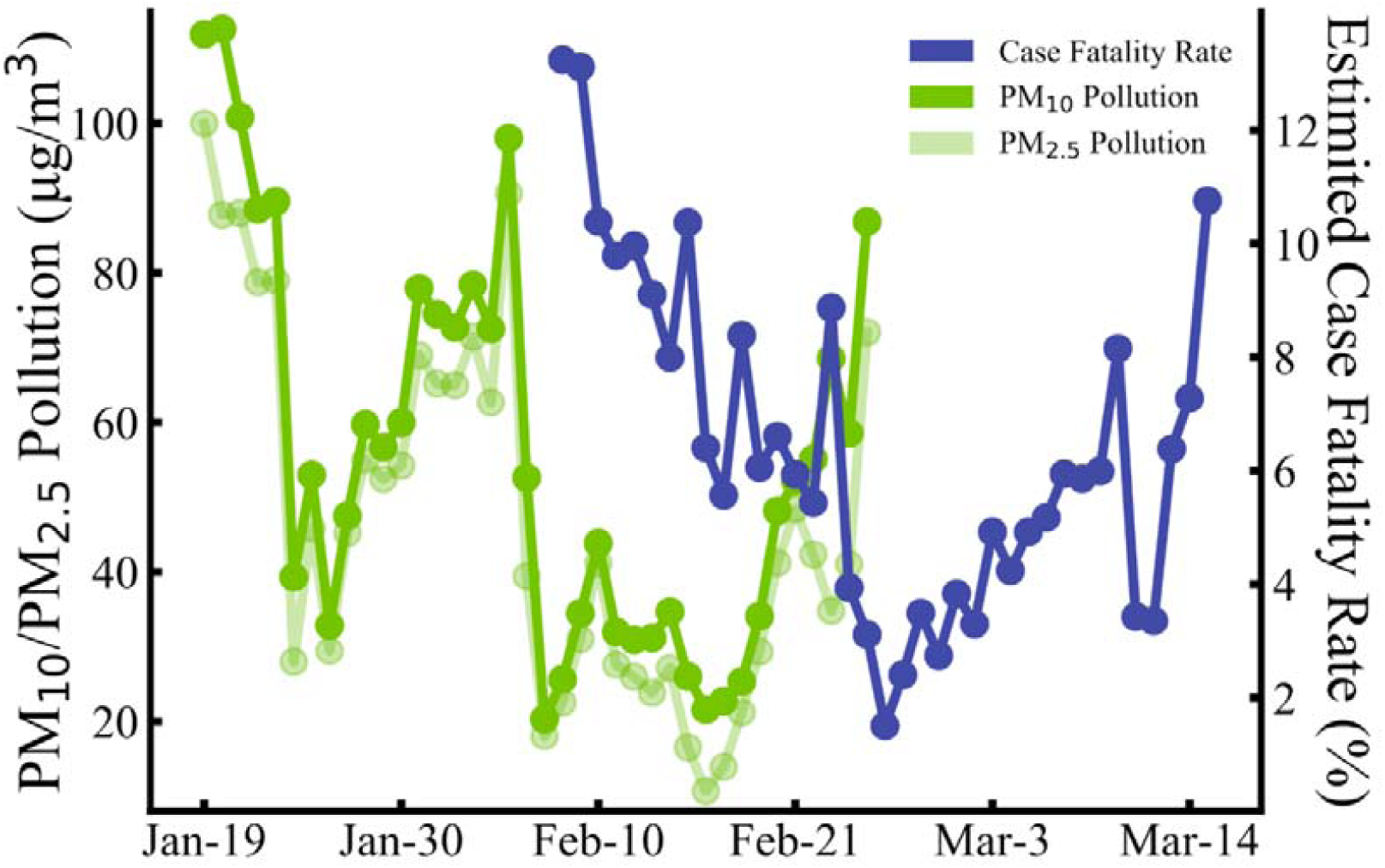
Daily Case Fatality Rate (blue points), PM_2.5_ (light green points) and PM_10_ (green points) Level from February 19 to March 15.

After adjustment for temperature and relative humidity, CFR was positively associated with all lag0 – lag5 concentrations of PM_2.5_ and PM_10_ (r>0.36, p<0.03), and the associations were the most significant with lag3 PM_2.5_ and PM_10_ (r=0.65, p=2.8×10^−5^ & r=0.66, p=1.9×10^−5^), suggesting that COVID-19 held higher case fatality rate with increasing concentrations of PM_2.5_ and PM_10_ in temporal scale (**Figure 2**). In addition, we did not find significance in the association between temperature or relative humanity with COVID-19 CFR (r=-0.13, p=0.44 & r=0.21, p=0.22).

**Figure 2.**
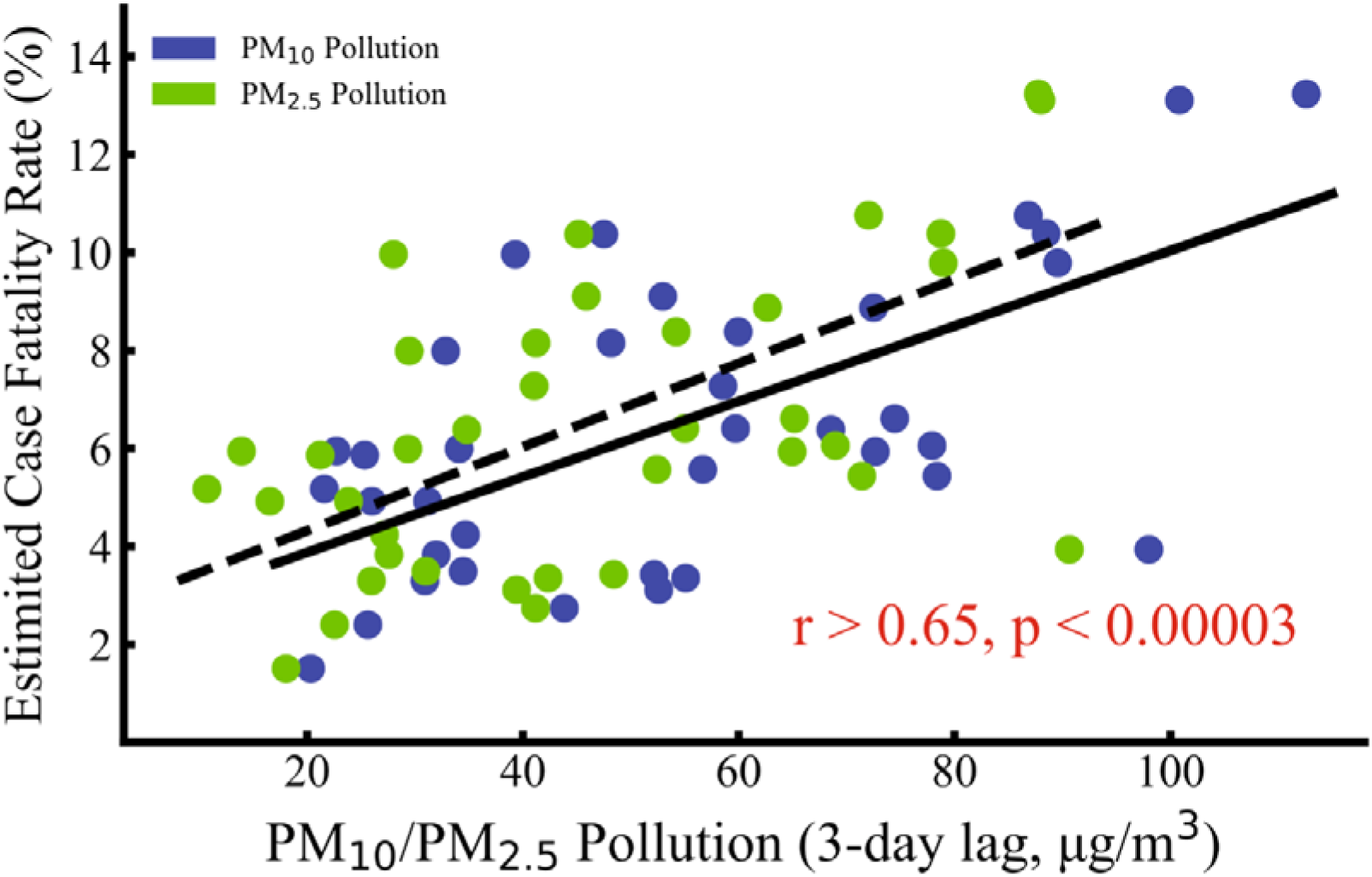
Daily Case Fatality Rate Versus PM_2.5_ and PM_10_ pollution. Case fatality rate was positively associated with 3-day lag PM_2.5_ (green points, r=0.65, p=2.8×10^−5^) and PM_10_ (blue points, r=0.66, p=1.9×10^−5^) pollution. Temperature and relative humidity effects have been removed during statistical analysis.

Moreover, PM_2.5_, PM_10_ and CFR significantly decreased over time between 19 January 2020 to 15 March 2020 (r=-0.34, p=0.038 & r=-0.45, p=0.0055 & r=-0.50, p=0.015), which may be affected by reduced human activities and improving medical support. After further adjustment for time effects, CFR of COVID-19 still held a strong positive association with concentrations of PM_2.5_ and PM_10_ (r=0.48, p=0.0043 & r=0.49, p=0.0027).

## Discussion

The results of this study indicated that the death of COVID-19 was highly correlated with PM_2.5_ and PM_10_ concentrations, which has been confirmed in other studies on respiratory diseases^6^. Most deaths of COVID-19 worldwide have been in older adults, especially those with underlying health problems, which made the population more vulnerability from air pollution. Considering the fact that the patients died from COVID-19 are likely to be critically ill, most of them might stay in ICU for treatment^7^. We speculated that the impact of PM_2.5_ and PM_10_ on death mainly affected the process of patients developed from mild to severe by potentially increasing system inflammation and oxidative stress, then decreasing the cardiopulmonary functions and finally influencing the prognosis of COVID-19 patients^8^. That might be the reason why only PM_2.5_ and PM_10_ of the first several days in the beginning of infections have significant associations with CFR.

The study was limited to a short period of season with less variation of air pollution. However, the correlation of trends of death and air pollution is quite convincing. In addition, there are also likely risks for the individuals with mild respiratory symptoms who are infected but never diagnosed, leading to a potential underestimated CFR. Longitudinal studies on a larger cohort would help to understand the accurate associations between CFR of COVID-19 and air pollution.

## Data Availability

We collected COVID-19 confirmed case information in China reported by the National Health Commission (http://www.nhc.gov.cn/xcs/xxgzbd/gzbd_index.shtml) and the Provincial Health Commissions of China (http://wjw.hubei.gov.cn/bmdt/ztzl/fkxxgzbdgrfyyq/);We also collected daily fine particulate matter (PM2.5), and inhalable particulate matter (PM10) from National Urban Air Quality Publishing Platform (http://106.37.208.233:20035/), and meteorological data including daily mean temperature and relative humidity from the China Meteorological Data Sharing Service System.

http://wjw.hubei.gov.cn/bmdt/ztzl/fkxxgzbdgrfyyq/

http://106.37.208.233:20035/

## Author contributions

Dr. Ye Yao, Jinhua Pan, Zhixi Liu and Xia Meng contributed equally.

Dr. Weibing Wang and Dr. Haidong Kan contributed equally.

Concept and design: Ye Yao, Haidong Kan, and Weibing Wang.

Acquisition, analysis, or interpretation of data: Ye Yao, Jinhua Pan, Zhixi Liu, Xia

Meng and Weidong Wang.

Drafting of the manuscript: Ye Yao, Jinhua Pan, Zhixi Liu.

Critical revision of the manuscript for important intellectual content: Haidong Kan, and Weibing Wang.

Statistical analysis: Ye Yao and Xia Meng.

## Acknowledgements

This study was sponsored by the Bill & Melinda Gates Foundation (Grant No. OPP1216424).

